# Effects of Statin Therapy in Patients Treated with Drug-Eluting and Drug-Coated Stents for Femoropopliteal Lesions: STAR-FP Study Outcomes

**DOI:** 10.1101/2024.09.06.24313216

**Authors:** Tatsuro Takei, Takahiro Tokuda, Naoki Yoshioka, Kenji Ogata, Akiko Tanaka, Shunsuke Kojima, Kohei Yamaguchi, Takashi Yanagiuchi, Tatsuya Nakama, the LEADers PAD investigators

**Author notes:** CORRESPONDENCE TO: Tatsuro Takei, MD 6-7 Izumi-cho, Kagoshima, Japan. Postal code 892-0821.

## Abstract

**Background:** The effects of statins on drug-eluting stents (DESs) and drug-coated stents (DCSs) for femoropopliteal (FP) lesions are not well known. Therefore, this multicenter retrospective evaluated the impact of statins on DES and DCS patency.

**Methods:** Between January 2018 and December 2021, 449 patients were treated with DES and DCS at eight cardiovascular centers in Japan (LEADers FP registry). These lesions were divided into statin-treated and non-statin-treated arms. After propensity score matching, the effects of statins on DES and DCS were evaluated. The 2-year primary outcome measure was stent patency. The secondary outcomes included secondary patency, clinically driven target lesion revascularization (CD-TLR), limb salvage, major adverse limb events (MALE, CD-TLR+ major amputation), and a composite of overall survival and MALE or all-cause death at 2 years.

**Results:** After propensity score matching, the baseline and procedural characteristics did not differ significantly between the 135 patient pairs in the statin and non-statin groups. The primary patency at two years was significantly better in the statin group than in the non-statin group (86.9% vs. 75.1%, p=0.041). Regarding the secondary endpoints, the statin group demonstrated significantly superior secondary patency and freedom from MALE or all-cause mortality (95.5% vs. 87.0%, p=0.023 and 73.7% vs. 60.0%, p=0.012, respectively).

**Conclusions:** The results of this retrospective multicenter study demonstrated the superior primary patency in the statin group compared with the non-statin group at two years. These findings suggest that statins improve patency in patients undergoing DES and DCS.

## What is known

- Statins are useful in the development and suppression of atherosclerotic diseases.
- Statins improve patency of bare-metal stents implanted for femoropopliteal (FP) lesions.

## What this study adds

- Statin use may improve the patency of drug-eluting and drug-coated stents for FP lesions.

## Background

The outcomes of endovascular therapy (EVT) using drug-eluting stents (DESs, ELUVIA, Boston Scientific, MA, USA) and drug-coated stents (DCSs, Zilver PTX, Cook Medical, Bloomington, IN) in femoropopliteal lesions have been reported and are widely applied in real-world practice.^1–3^ The drugs applied to these stents improve patency compared with bare nitinol stents. However, restenosis has not been completely resolved.^4,5^ While statins reportedly improve the patency of stents implanted in femoropopliteal lesions,^6–10^ previous studies assessed bare nitinol stents, which differ from the current situation in which paclitaxel devices, such as DES and DCS, are used. These studies were also limited by their small sample sizes, single-center analysis, and lack of propensity score matching. In addition to reducing low-density lipoprotein (LDL) cholesterol levels, statins reportedly to improve the prognosis of patients with coronary artery disease and lower extremity arterial disease.^11,12^ Statins also relieve ischemic symptoms in the lower extremities.^13^ In the present study, we retrospectively analyzed multicenter real-world data on femoropopliteal lesions in which DES or DCS was implanted to assess the clinical impact of statins. The main objective was to investigate the effect of DES with or without statins on DES and DCS patency rates. Also, as described above, as statins have a variety of effects, the secondary endpoints included limb and life expectancies to assess the impact of statins.

## Materials and Methods

### Ethical considerations

This study was approved by the medical ethics committees of the investigators’ respective hospitals and was conducted in accordance with the principles of the Declaration of Helsinki. Because this was a retrospective observational study that did not require intervention, the requirement for written informed consent was waived.

Consent for publication in this retrospective study was obtained from the enrolled patients before the interventional procedure.

### Patient population

The LEADers registry enrolled 2173 patients who underwent EVT for de novo femoropopliteal lesions between January 2018 and December 2021 at eight Japanese institutions (Sendai Kosei Hospital, Tokyo Bay Medical Center, Saiseikai Yokohama Eastern Hospital, Nagoya Heart Center, Ogaki Municipal Hospital, Miyazaki Medical Association Hospital, Rakuwakai Otowa Hospital, and Tenyokai Central Hospital). The Effects of **S**tatin **T**herapy in P**A**tients Treated with D**R**ug-Eluting and Drug-Coated Stents for **F**emoro**P**opliteal Lesions (STAR-FP) study retrospectively analyzed data from 449 patients from the registry who underwent implantation of an ELUVIA stent or Zilver PTX for femoropopliteal lesions. All enrolled patients had intermittent claudication, rest pain, or ischemic foot wounds (Rutherford classification 1–6). This cohort also included patients treated with ELUVIA or Zilver PTX combined with other devices (drug-coated balloons, bare nitinol stents, and stent-grafts). The patients were divided into statin and non-statin groups based on their statin therapy status at the time of EVT. The first lesion was used as the reference lesion for patients whose bilateral limbs were treated. In addition, propensity-matched analyses were performed to adjust for patient and lesion characteristics and background.

Independent investigators collected patient-specific, angiographic, procedural, and follow-up data from individual hospital databases. Follow-up after EVT was performed according to the protocols of the individual hospitals and physicians and included ankle-brachial index (ABI), duplex ultrasound, or lower-extremity angiography.

### Interventional procedures

Interventional procedures were performed with a 5-7 Fr sheath inserted into the common femoral artery via a crossover or ipsilateral approach. Immediately before the procedure, 5,000 U of heparin was administered intra-arterially. Guidewires with diameters of 0.014, 0.018, and 0.035 inches were used. The treatment strategy was determined at the discretion of each operator. The need for antithrombotic therapy or statin administration was determined by the respective physicians and operators.

### Outcome assessments

Restenosis was defined as a peak systolic velocity ratio of ≧2.4 by duplex ultrasound, a decrease in ABI of 0.15 or more, and ≧50% stenosis or obstruction on lower extremity angiography. Target lesion revascularization (TLR) was defined as the need for reintervention or surgical revascularization.

Primary patency was defined as the absence of restenosis or recanalization of the treated lesion. Secondary patency was defined as the absence of re-occlusion.

Major adverse limb events (MALEs) included a composite of clinically driven TLR (CD-TLR) and major amputations.

### Statistical analysis

The statistical analyses were performed using JMP Pro version 16 (SAS Institute, Inc., Cary, NC, USA). SAS version 9.4 (SAS Institute, Inc., Cary, NC, USA) was used to perform the interaction analyses. Data on the baseline patient and lesion characteristics are presented as means ± standard deviation for continuous variables and as frequencies (percentages) for categorical variables, unless otherwise noted.

Only one dataset was represented by median values, as noted. Statistical significance was set at p<0.05. Differences in baseline characteristic categories between the groups were analyzed using t-tests and chi-square tests for continuous and categorical variables, respectively. When comparing clinical outcomes between the statin and non-statin groups, propensity score matching was used to minimize the effect of group differences on baseline characteristics. The propensity scores were calculated using a logistic regression model. Logistic analysis was performed and matched according to age, sex, symptoms, body mass index, ambulatory status, smoking, hypertension, diabetes, chronic kidney disease, medications, history of coronary artery disease, ankle-brachial pressure ratio, lesion location, reference vessel diameter, lesion length, chronic total occlusion (CTO), peripheral artery calcium scoring system classification, and below-knee outflow. A caliper cutoff of 0.2 was used for propensity score matching.

## Results

### Baseline characteristics

As shown in the study flowchart (Figure 1), 135 pairs (statin and non-statin groups) were evaluated for clinical outcomes after propensity score matching. Baseline characteristics of the patients are shown in Table 1. The overall population (n=449) demonstrated significant differences between the statin (n=264) and non-statin groups (n=185) in terms of age, ambulatory status, diabetes, coronary artery disease, dialysis, symptoms, ABI, and lesion length. Baselines between the two matched groups were analyzed, and all p-values were >0.05. Regarding the standardized differences, the CTO length was only slightly longer in the statin group.

**Figure 1.**
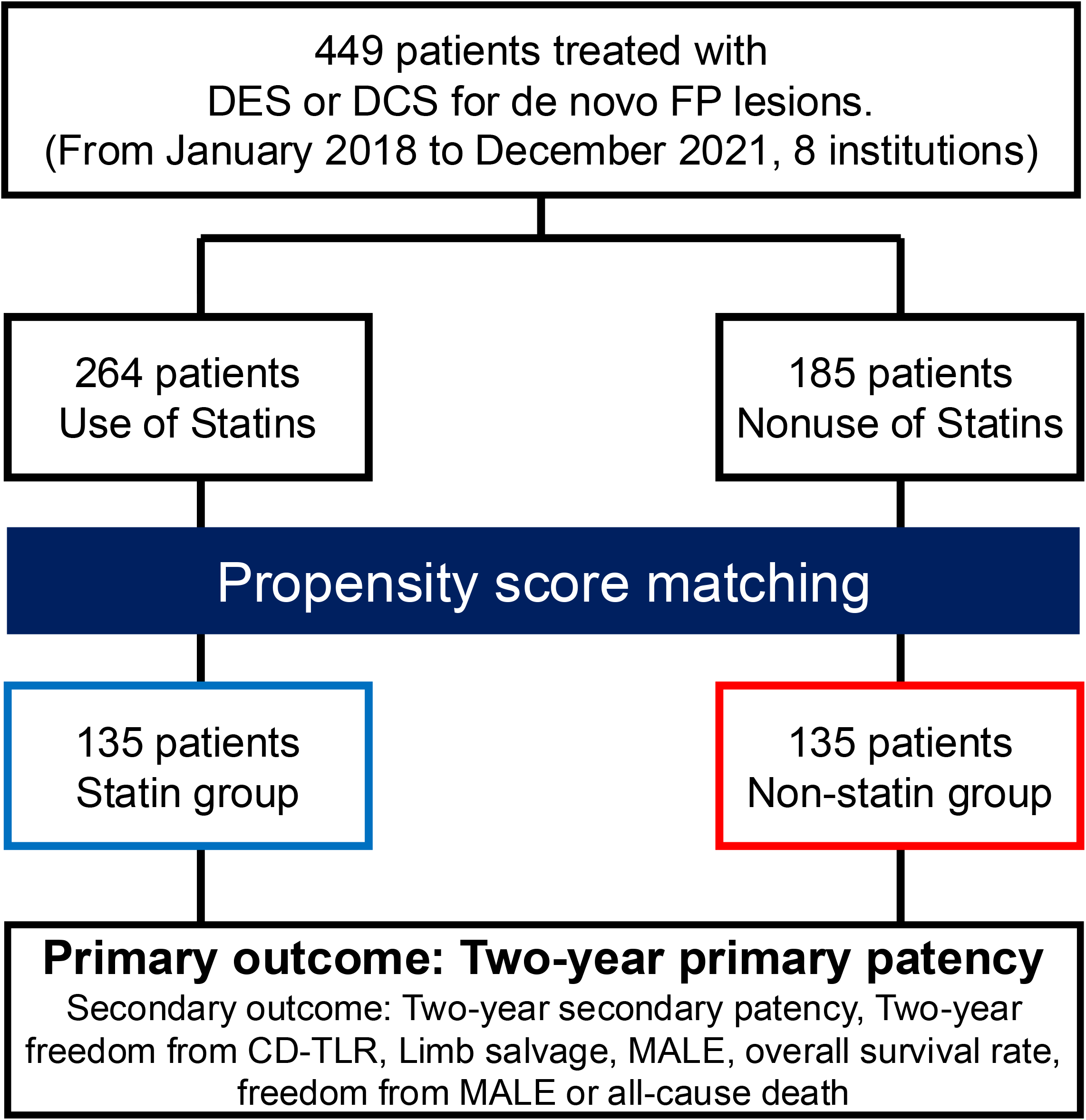
Study flowchart. DES, drug-eluting stent; DCS, drug-coated stent; FP, femoropopliteal; CD-TLR, clinically driven target lesion revascularization; MALE, major adverse limb event

**Table 1.**
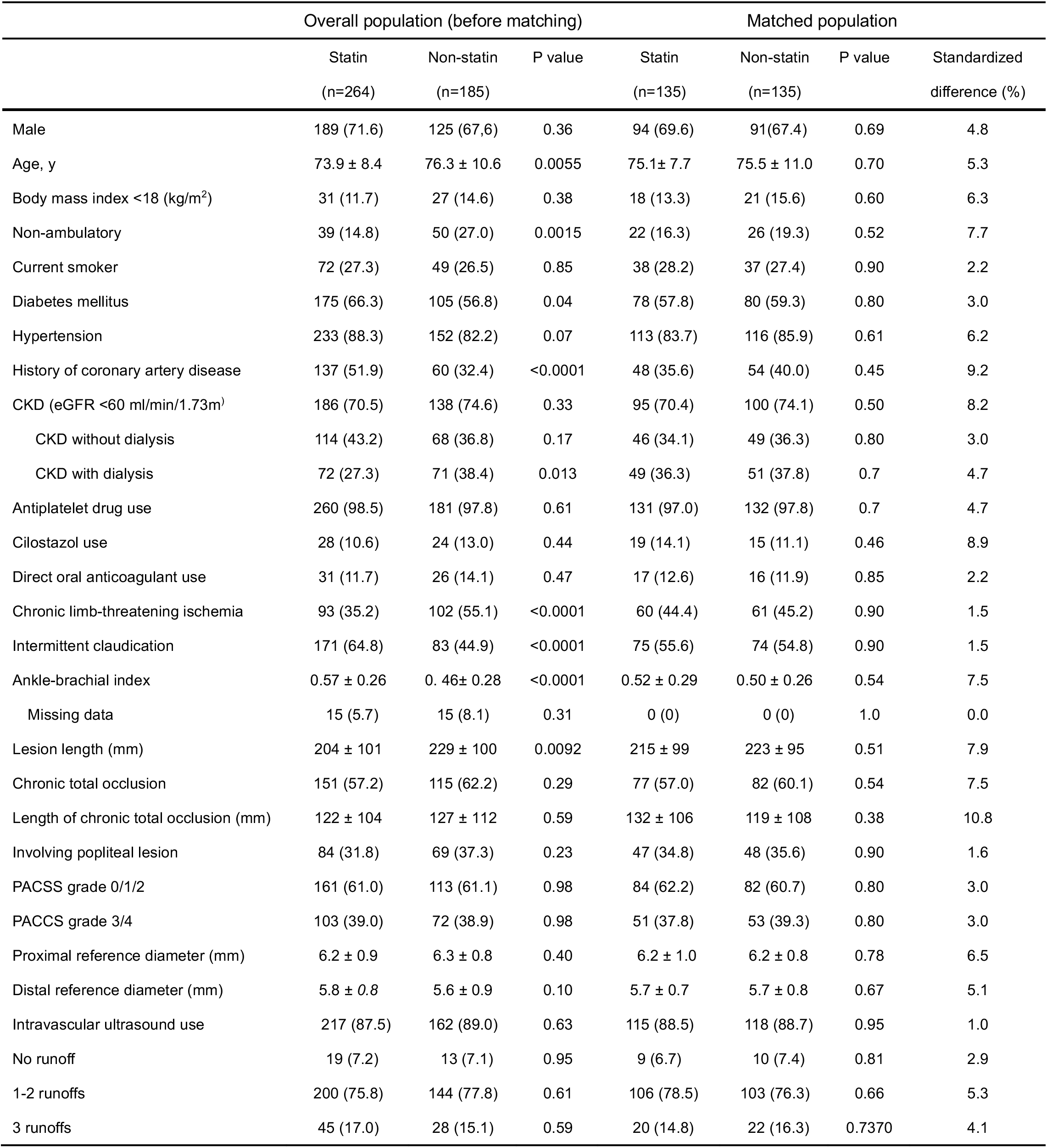
Baseline characteristics before and after matching. Values are expressed as means ± standard deviations or numbers (percentages). CKD, chronic kidney disease; PACSS, peripheral artery calcium scoring

### Outcome measures

Table 2 shows cholesterol levels and procedural characteristics in the matched population. Total and LDL cholesterol levels differed significantly between the two groups (159 ± 37 mg/dl vs. 174 ± 41 mg/dl, p=0.0027, and 84 ± 31 mg/dl vs. 99 ± 31 mg/dl, p=0.0002, respectively). High-density lipoprotein (HDL) levels did not differ significantly between the groups. In the statin group, 95.6% and 4.4% of patients were on strong and standard statins, respectively. The use of ELUVIA and Zilver PTX and drug-coated balloons or other scaffold devices in combination therapy did not differ significantly between the two groups. Regarding drug-coated balloon types, the Ranger (Boston Scientific, MA, USA) was more frequently used in the non-statin group (0% vs. 2.2%, p=0.04, respectively), while the IN.PACT (Medtronic, MN, USA) and LUTONIX (BD, NJ, USA) were statistically equivalent (9.6% vs. 14.1%, p=0.25, 7.4% vs. 3.0%, p=0.81, respectively). Regarding scaffold devices, the use of bare nitinol stents and VIABAHN (Gore, Flagstaff, AZ) did not differ significantly between the two groups. The nitinol stents used in this study were the S.M.A.R.T. (Cordis Corporation, Miami Lakes, Florida) and Supera (Abbott Vascular, Santa Clara, CA). Residual stenosis, stent size diameter, and stent length also did not differ significantly between the two groups.

**Table 2.** Cholesterol levels and procedural characteristics after matching. Values are expressed as means ± standard deviations or numbers (percentages). LDL, low-density lipoprotein; HDL, high-density lipoprotein; DCB, drug-coated balloon.

|  | Statin<br>(n=135) | Non-statin<br>(n=135) | P value |
| --- | --- | --- | --- |
| Total cholesterol (mg/dl) | 159 ± 37 | 174 ± 41 | 0.0027 |
| Missing data | 3 (2.2) | 6 (4.5) | 0.30 |
| LDL cholesterol (mg/dl) | 84 ± 31 | 99 ± 31 | 0.0002 |
| Missing data | 2 (1.5) | 5 (3.7) | 0.20 |
| HDL cholesterol (mg/dl) | 48 ± 15 | 49 ± 19 | 0.77 |
| Missing data | 2 (1.5) | 6 (4.5) | 0.14 |
| Strong statin | 129 (95.6) | - | - |
| Standard statin | 6 (4.4) | - | - |
| Zilver PTX stent | 14 (10.4) | 10 (7.4) | 0.39 |
| ELUVIA stent | 121 (89.6) | 126 (93.3) | 0.27 |
| Combination with DCB | 23 (17.0) | 31 (23.0) | 0.22 |
| Use of IN.PACT | 13 (9.6) | 19 (14.1) | 0.25 |
| Use of Ranger | 0 (0) | 3 (2.2) | 0.04 |
| Use of Lutonix | 10 (7.4) | 4 (3.0) | 0.81 |
| Combination with other scaffold device | 7 (5.2) | 4 (3.0) | 0.35 |
| Use of Bare nitinol stent (Supera or S.M.A.R.T.) | 5 (3.7) | 2 (1.5) | 0.24 |
| Use of VIABAHN | 2 (1.5) | 2 (1.5) | 1.0 |
| Residual stenosis after stent implantation | 0 (0, 50) | 0 (0, 40) | 0.74 |
| Stent size (mm) | 6.4 ± 0.5 | 6.4 ± 0.5 | 0.91 |
| Stent length (mm) | 219 ± 102 | 210 ± 102 | 0.49 |

Values are expressed as the means ± standard deviations or numbers (percentages). The residual stenosis after stent implantation was expressed as median (minimum, maximum). LDL, low density lipoprotein; HDL, high density lipoprotein, DCB, drug coating balloon.

Figure 2 shows the 2-year primary patency rates for the statin and non-statin groups. Primary patency was significantly higher in the statin group than in the non-statin group (86.9% vs. 75.1%, p=0.041). The mean follow-up period was 460 ± 265 days, and 36 patients in the matched population developed restenosis at the treated site.

**Figure 2.**
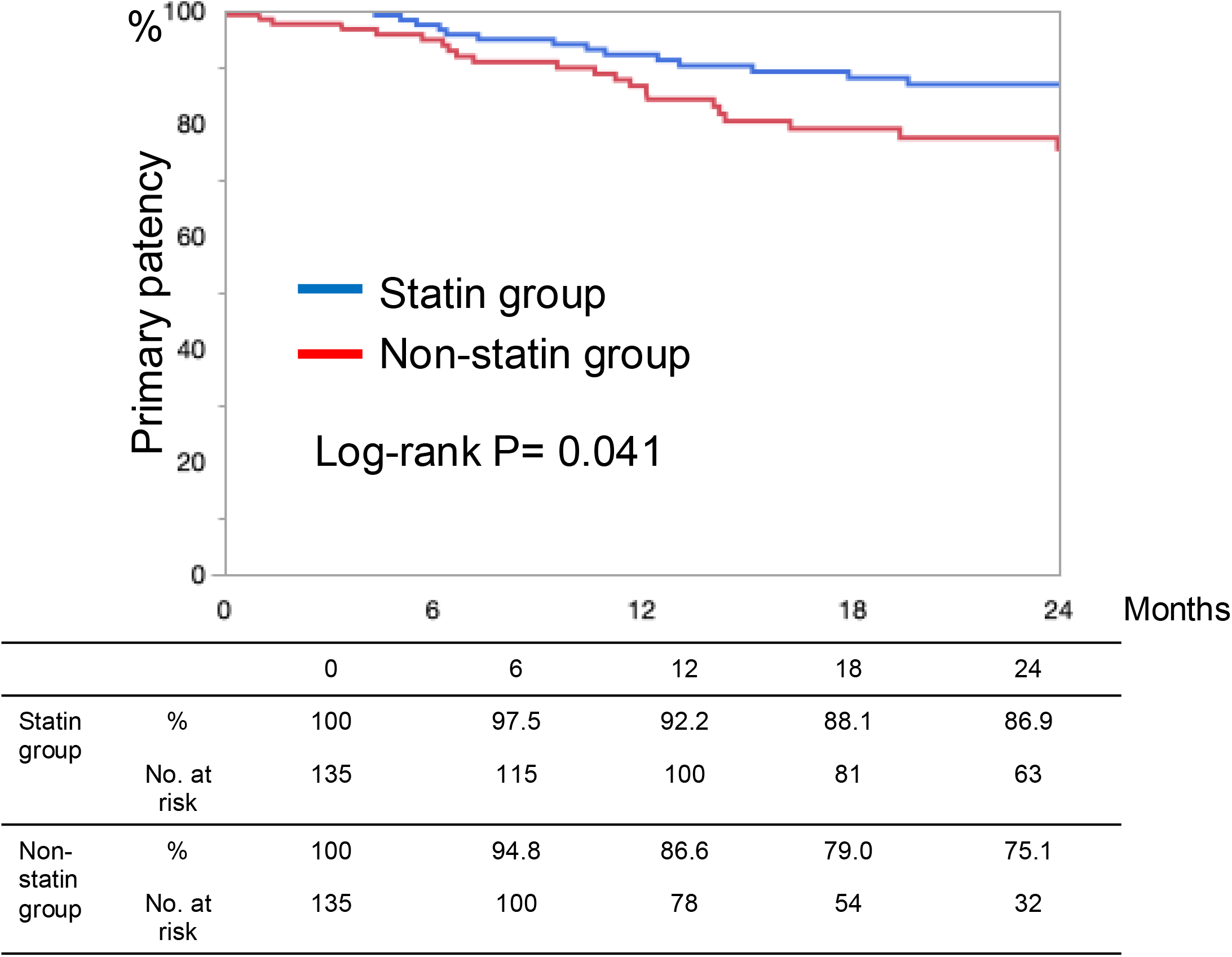
Primary patency in the matched population.

Figure 3 shows each clinical outcome, including the 2-year secondary patency. The two-year secondary patency rate was significantly higher in the statin group than in the non-statin group (95.5% vs. 87.0%, p=0.023). However, freedom from CD-TLR, limb salvage, freedom from MALE, and overall survival rates did not differ significantly between the two groups (85.5% vs. 84.2%, p=0.37; 97.4% vs. 95.5%, p=0.42; 86.9% vs. 79.9%, p=0.17; and 80.9% vs. 72.3%, p=0.07, respectively). One of the composite outcomes, freedom from MALE or all-cause death, was significantly superior in the statin group than in the non-statin group (73.7% vs. 60.0%, p=0.012). Evaluation of freedom from acute limb ischemia due to stent thrombosis using Kaplan–Meier estimates showed no significant difference between the two groups (98.4% vs. 97.5%, p=0.62).

**Figure 3.**
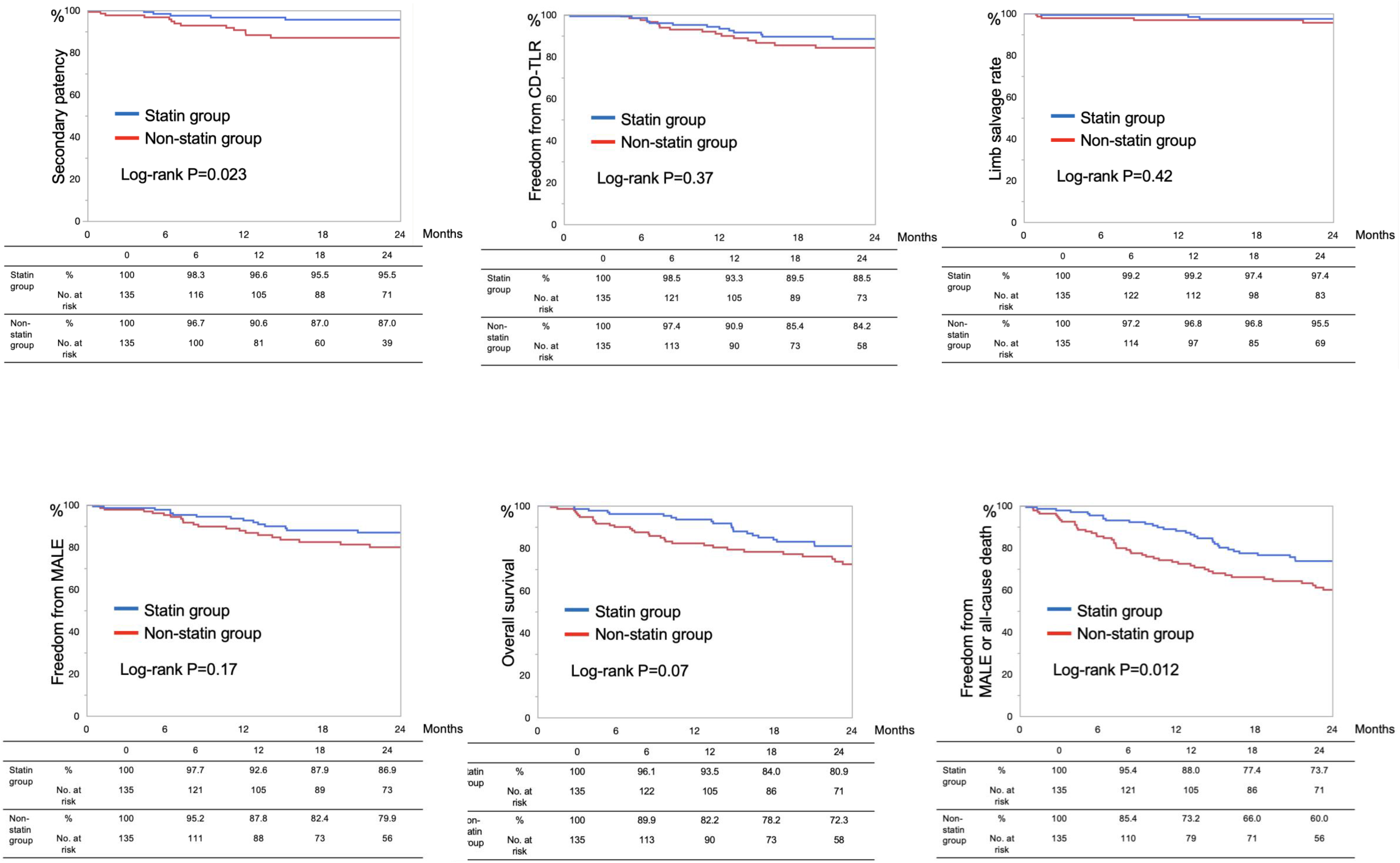
Secondary outcome.

Interactions in each subgroup with respect to primary patency are shown in Figure 4. No interaction was observed in any of the subgroups (p≥0.05). Regarding sex, male patients tended to benefit more from statin therapy compared with female patients (p=0.06).

**Figure 4.**
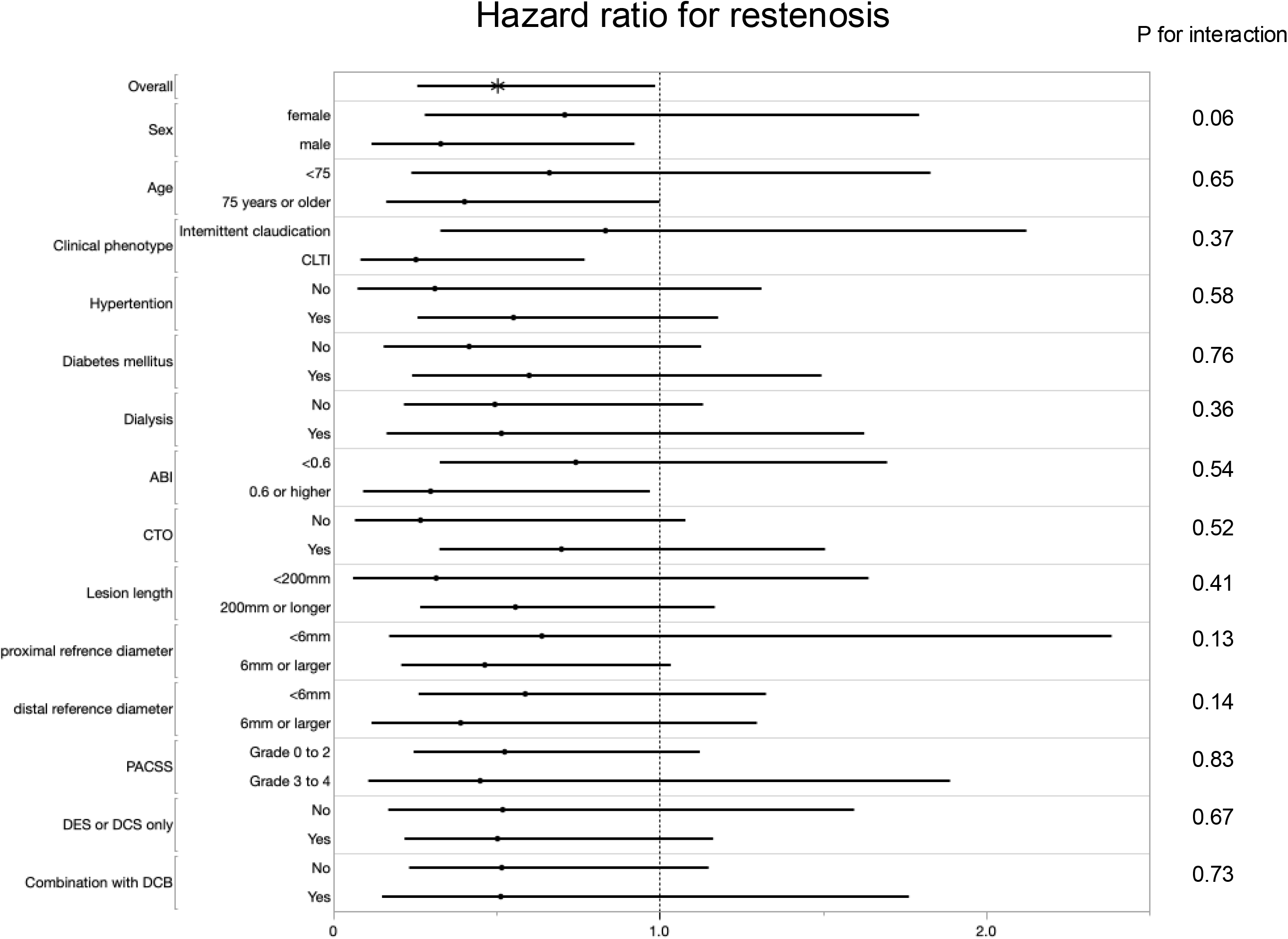
Interaction analysis for each subgroup with respect to primary patency CLTI, chromic limb-threatening ischemia; ABI, ankle-brachial index; CTO, chronic total occlusion; PACSS, peripheral artery calcium scoring system; DES, drug-eluting stent; DCS, drug-coated stent; DCB, drug-coated balloon.

## Discussion

In this study, based on LEADers registry data, the statin group showed significantly better primary patency compared with the non-statin group. The Kaplan–Meier curves for primary and secondary patency showed differences between the two groups starting around 12 and 6 months, respectively. This finding may be attributed to the effect of statins in preventing atherosclerosis and the progression of in-stent restenosis.^14,15^ In addition to lowering LDL cholesterol levels and inhibiting atherosclerosis progression, statins also have anti-inflammatory and endothelial function-improving effects, which may improve the patency rates of DES and DCS.^16^ However, statins are not effective in preventing perioperative patency loss in bypass grafts after DES or DCS implantation.^17^ Unfortunately, the results of the present study also showed no effect of statins on improving freedom from CD-TLR, limb salvage, or MALE.

The statin group in the present study demonstrated superior primary and secondary patency rates compared with the non-statin group. The incidence of stent occlusion is relatively high among recurrent ELUVIA-treated lesions.^18^ Moreover, the incidence of cumulative stent occlusion does not plateau and continues to increase in Zilver PTX.^19^ As completely occluded stent restenosis is associated with increased risks of recurrent refractory stent restenosis and surgical revascularization, the clinical utility of statins, which may improve secondary patency, may be significant.^20^ The results of a study examining the mechanism of in-stent reocclusion using angioscopy suggested the potential influence of organic stenosis in the stent or at the edge.^21^ Therefore, the better secondary patency in the statin group in the present study may have been due to the inhibition of organic stenosis progression.

European, United States, and Japanese guidelines strongly recommend the administration of statins to patients with lower extremity arterial disease (LEAD) to improve life expectancy.^22,23^ In the present study, the administration of statins to only 58.8% of patients may have resulted in an underestimation of LEAD. The log-rank test results of the 2-year overall survival rate in this study showed a better trend in the statin group (p=0.07). The results of the Wilcoxon test showed a significantly better prognosis in the statin group than in the non-statin group (p=0.036).

Evaluation of the composite of MALE or all-cause death, which was assessed as the best endovascular versus surgical therapy in patients with chronic limb-threatening ischemia.^24^ showed that freedom from MALE or all-cause death was significantly higher in the statin group than in the non-statin group, suggesting the efficacy of statins.

The results of the interaction analysis showed no significant differences between the subgroups (p>0.05). Although not statistically significant, a trend toward an interaction between sexes was observed. A meta-analysis on the effects of statins reported no significant differences between men and women,^25^ possibly due to the high proportion of men (approximately 70%) and low number of women in this cohort.

In addition to the aforementioned reports of statins improving patency in the femoropopliteal region, statins also reportedly improve patency in iliac artery stents and bypass grafts.^26–28^ Future studies should examine the effects and long-term outcomes of statin use in other cohorts.

## Limitations

This study has several limitations. For instance, while information on statin dosing was available at the time of EVT, compliance with subsequent dosing was unknown. Additionally, the LEADers registry is a retrospective study and it is difficult to assess compliance with statin medications. This study also performed propensity score matching using lesion length and CTO presence or absence; however, the standardized difference was slightly >10% for CTO lesion length (p=0.38). Stuart reported that a standardized difference of <25% is a balanced matching model; thus, a value of 10.8 should not significantly affect the results.^29^ As this study used real-world data, it included cases in which the device was used in combination with other devices. Therefore, an interaction analysis was performed, which showed no interaction in the subgroups treated with DCB plus DES or DCS versus DCS or DES alone.

## Conclusion

The results of this retrospective multicenter study demonstrated superior primary and secondary patency in the statin group compared with the non-statin group at 2 years. These findings suggest that statins may be effective in improving DES and DCS patency. Freedom from a composite of MALE and all-cause death at 2 years was also higher in the statin group than in the non-statin group.

## Availability of data and materials

The datasets used and/or analyzed in this study are available from the corresponding author upon reasonable request.

## Non-standard Abbreviations and Acronyms

EVT: endovascular therapy
DES: drug-eluting stent
DCS: drug-coated stent
FP: femoropopliteal
CD-TLR: clinically driven-target lesion revascularization
MALE: major adverse limb event
CTO: chronic total occlusion
LEAD: lower extremity arterial disease

## Acknowledgments

We thank Editage for editing and reviewing the manuscript for English language.

## Consent for publication

## References

1. Iida O, Fujihara M, Kawasaki D, Mori S, Yokoi H, Miyamoto A, Kichikawa K, Nakamura M, Ohki T, Diaz-Cartelle J, et al. 24-Month efficacy and safety results from Japanese patients in the IMPERIAL randomized study of the Eluvia drug- eluting stent and the Zilver PTX drug-coated stent. Cardiovasc Intervent Radiol 2021;44:1367–1374. doi: 10.1007/s00270-021-02901-6.

2. Müller-Hülsbeck S, Benko A, Soga Y, Fujihara M, Iida O, Babaev A, O’Connor D, Zeller T, Dulas DD, et al. Two-Year Efficacy and Safety Results from the IMPERIAL randomized study of the Eluvia polymer-coated drug-eluting stent and the Zilver PTX polymer-free drug-coated stent. Cardiovasc Intervent Radiol. 2021;44:368–375. doi: 10.1007/s00270-020-02693-1.

3. Shibata T, Iba Y, Shingaki M, Yamashita O, Tsubakimoto Y, Kimura F, Hatada A, Kasashima F, Ueno K, Nakanishi K, et al. One year outcomes of Zilver PTX versus Eluvia for femoropopliteal disease in real-world practice: REALDES Study. J Endovasc Ther. 2023:15266028231179861. doi: 10.1177/15266028231179861.

4. Kurata N, Iida O, Takahara M, Asai M, Okamoto S, Ishihara T, Nanto K, Tsujimura T, Hata Y, Toyoshima T, et al. Comparing predictors influencing restenosis following high-dose drug-coated balloon angioplasty and fluoropolymer-based drug-eluting stenting in femoropopliteal artery lesions. J Endovasc Ther. 2023:15266028231209234. doi:10.1177/15266028231209234.

5. Soga Y, Fujihara M, Tomoi Y, Iida O, Ishihara T, Kawasaki D, Ando K. One-year late lumen loss between a polymer-coated paclitaxel-eluting stent (Eluvia) and a polymer-free paclitaxel-coated stent (Zilver PTX) for femoropopliteal disease. J Atheroscler Thromb. 2020:164–171. doi: 10.5551/jat.50369.

6. Kim W, Gandhi RT, Peña CS, Herrera RE, Schernthaner MB, Acuña JM, Becerra VN, Katzen BT. The influence of statin therapy on restenosis in patients who underwent nitinol stent implantation for de novo femoropopliteal artery disease: two-year follow-up at a single center. J Vasc Interv Radiol. 2016:1494–501. doi: 10.1016/j.jvir.2016.05.037.

7. de Grijs D, Teixeira P, Katz S. The association of statin therapy with the primary patency of femoral and popliteal artery stents. J Vasc Surg 2018:1472–1479. doi: 10.1016/j.jvs.2017.09.022.

8. Siracuse JJ, Gill HL, Cassidy SP, Messina MD, Catz D, Egorova N, Parrack I, McKinsey JF. Endovascular treatment of lesions in the below-knee popliteal artery. J Vasc Surg. 2014:356–361. doi: 10.1016/j.jvs.2014.02.012. Epub 2014 Mar 18.

9. Aiello FA, Khan AA, Meltzer AJ, Gallagher KA, McKinsey JF, Schneider DB. Statin therapy is associated with superior clinical outcomes after endovascular treatment of critical limb ischemia. J Vasc Surg. 2012:371–379; discussion 380. doi: 10.1016/j.jvs.2011.08.044.

10. Braun SK, Jorge DW, Bortolanza G, da Rocha JBT. Effects of statin use on primary patency, mortality, and limb loss in patients undergoing lower-limb arterial angioplasty: a systematic review and meta-analysis. Int J Clin Pharm. 2023:17– 25. doi: 10.1007/s11096-022-01513-5.

11. Westin GG, Armstrong EJ, Bang H, Yeo KK, Anderson D, Dawson DL, Pevec WC, Amsterdam EA, Laird JR. Association between statin medications and mortality, major adverse cardiovascular event, and amputation-free survival in patients with critical limb ischemia. J Am Coll Cardiol 2014:682–690. doi: 10.1016/j.jacc.2013.09.073.

12. Orkaby AR, Driver JA, Ho YL, Lu B, Costa L, Honerlaw J, Galloway A, Vassy JL, Forman DE, Gaziano JM, et al. Association of statin use with all-cause and cardiovascular mortality in US veterans 75 years and older. JAMA. 2020:68–78. doi: 10.1001/jama.2020.7848. Erratum in: JAMA. 2020;3241468.

13. Mohler ER 3rd, Hiatt WR, Creager MA. Cholesterol reduction with atorvastatin improves walking distance in patients with peripheral arterial disease. Circulation. 2003 (12):1481–1486. doi: 10.1161/01.CIR.0000090686.57897.F5.

14. Watts GF, Lewis B, Brunt JN, Lewis ES, Coltart DJ, Smith LD, Mann JI, Swan AV. Effects on coronary artery disease of lipid-lowering diet, or diet plus cholestyramine, in the St Thomas’ Atherosclerosis Regression Study (STARS). Lancet. 1992;339:563–599. doi: 10.1016/0140-6736(92)90863-x.

15. Walter DH, Schächinger V, Elsner M, Mach S, Auch-Schwelk W, Zeiher AM. Effect of statin therapy on restenosis after coronary stent implantation. Am J Cardiol. 2000;85:962–968. doi: 10.1016/s0002-9149(99)00910-8

16. Momin A, Melikian N, Wheatcroft SB, Grieve D, John LC, El Gamel A, Marrinan MT, Desai JB, Driver C, Sherwood R, et al. The association between saphenous vein endothelial function, systemic inflammation, and statin therapy in patients undergoing coronary artery bypass surgery. J Thorac Cardiovasc Surg. 2007;134:335–341. doi: 10.1016/j.jtcvs.2006.12.064.

17. Scali ST, Beck AW, Nolan BW, Stone DH, De Martino RR, Chang CK, Rzucidlo EM, Walsh DB. Completion duplex ultrasound predicts early graft thrombosis after crural bypass in patients with critical limb ischemia. J Vasc Surg. 2011;54:1006–1110. doi: 10.1016/j.jvs.2011.04.021.

18. Iida O, Takahara M, Soga Y, Yamaoka T, Fujihara M, Kawasaki D, Ichihashi S, Kozuki A, Nanto S, Sakata Y, et al. 1-Year outcomes of fluoropolymer-based drug-eluting stent in femoropopliteal practice: predictors of restenosis and aneurysmal degeneration. JACC Cardiovasc Interv. 2022;15:630–638. doi: 10.1016/j.jcin.2022.01.019.

19. Iida O, Takahara M, Soga Y, Nakano M, Yamauchi Y, Zen K, Kawasaki D, Nanto S, Yokoi H, Uematsu M, et al. 1-Year results of the ZEPHYR Registry (Zilver PTX for the femoral artery and proximal popliteal artery): Predictors of Restenosis. JACC Cardiovasc Interv. 2015;8:1105–1112.

20. Tosaka A, Soga Y, Iida O, Ishihara T, Hirano K, Suzuki K, Yokoi H, Nanto S, Nobuyoshi Ml. Classification and clinical impact of restenosis after femoropopliteal stenting. J Am Coll Cardiol. 2012 3;59:16–23. doi: 10.1016/j.jacc.2011.09.036.

21. Ishihara T, Iida O, Okamoto S, Fujita M, Masuda M, Nanto K, Shiraki T, Kanda T, Tsujimura T, Okuno S, et al. Potential mechanisms of in-stent occlusion in the femoropopliteal artery: an angioscopic assessment. Cardiovasc Interv Ther. 2017;32:313–317. doi: 10.1007/s12928-016-0411-3.

22. Aboyans V, Ricco JB, Bartelink MEL, Björck M, Brodmann M, Cohnert T, Collet JP, Czerny M, De Carlo M, Debus S, et al. 2017 ESC Guidelines on the Diagnosis and Treatment of Peripheral Arterial Diseases, in collaboration with the European Society for Vascular Surgery (ESVS): Document covering atherosclerotic disease of extracranial carotid and vertebral, mesenteric, renal, upper and lower extremity arteries Endorsed by: the European Stroke Organization (ESO), The Task Force for the Diagnosis and Treatment of Peripheral Arterial Diseases of the European Society of Cardiology (ESC) and of the European Society for Vascular Surgery (ESVS). Eur Heart J. 201839:763– 816. doi: 10.1093/eurheartj/ehx095.

23. Gerhard-Herman MD, Gornik HL, Barrett C, Barshes NR, Corriere MA, Drachman DE, Fleisher LA, Fowkes FG, Hamburg NM, Kinlay S, et al. 2016 AHA/ACC Guideline on the Management of Patients With Lower Extremity Peripheral Artery Disease: Executive Summary: A Report of the American College of Cardiology/American Heart Association Task Force on Clinical Practice Guidelines. J Am Coll Cardiol. 2017;69:1465–1508. doi: 10.1016/j.jacc.2016.11.008. Epub 2016 Nov 13. Erratum in: J Am Coll Cardiol. 2017;69:1520. doi: 10.1016/j.jacc.2017.02.003.

24. Farber A, Menard MT, Conte MS, Kaufman JA, Powell RJ, Choudhry NK, Hamza TH, Assmann SF, Creager MA, Cziraky MJ, et al. Surgery or endovascular therapy for chronic limb-threatening ischemia. N Engl J Med. 2022;387:2305– 2316. doi: 10.1056/NEJMoa2207899. Epub 2022 Nov 7.

25. Kostis WJ, Cheng JQ, Dobrzynski JM, Cabrera J, Kostis JB. Meta-analysis of statin effects in women versus men. J Am Coll Cardiol. 2012;59:572–582. doi: 10.1016/j.jacc.2011.09.067. Erratum in: J Am Coll Cardiol. 2012 Apr 17;59(16):1491.

26. Haraguchi T, Masanaga T, Fujita T, Otake R, Hachinohe D, Kaneko U, Kashima Y, Sato K. Comparative 2-year outcomes of the Misago stent versus other self- expandable nitinol stents for the endovascular treatment of aortoiliac disease. J Cardiovasc Surg (Torino). 2023;64:422–429. doi: 10.23736/S0021-9509.23.12500-6.

27. Henke PK, Blackburn S, Proctor MC, Stevens J, Mukherjee D, Rajagopalin S, Upchurch GR Jr, Stanley JC, Eagle KA. Patients undergoing infrainguinal bypass to treat atherosclerotic vascular disease are underprescribed cardioprotective medications: effect on graft patency, limb salvage, and mortality. J Vasc Surg. 200439:357–65. doi: 10.1016/j.jvs.2003.08.030.

28. Klingelhoefer E, Bergert H, Kersting S, Ludwig S, Weiss N, Schönleben F, Grützmann R, Gäbel G. Predictive factors for better bypass patency and limb salvage after prosthetic above-knee bypass reconstruction. J Vasc Surg. 2016 Aug;64(2):380–388.e1. doi: 10.1016/j.jvs.2016.02.059.

29. Stuart EA. Matching methods for causal inference: A review and a look forward. Stat Sci. 2010;25:1–21. doi: 10.1214/09-STS313.

